# The Protection Motivation Theory as an explanatory model for Intention to use Alcohol Protective Behavioral Strategies among Young Adults

**DOI:** 10.1101/2024.01.26.24301710

**Authors:** Bella M. González-Ponce, José Carmona-Márquez, Angelina Pilatti, Carmen Díaz-Batanero, Fermín Fernández-Calderón

## Abstract

**Aims:** This study aimed to prospectively examine the explanatory value of the Protection Motivation Theory (PMT) for the intention to use MD PBS, and to explore its invariance across genders

**Method:** A targeted sampling procedure was used to recruit 339 young adults in the community (*M_ean age_* = 21.1; *SD* = 2.21; female = 50.7%) who completed baseline and 2-month follow-up measures of the constructs of the PMT and for intentions to use each of the five MD PBS.

**Results:** The results of regression analyses revealed that the coping appraisal components (response efficacy and self-efficacy) had greater explanatory power for intention to use PBS than the threat appraisal components (perceived vulnerability and perceived severity). Perceived vulnerability to alcohol consequences was not prospectively associated with any specific behavioral intention or with the total MD PBS score. In contrast, perceived severity was prospectively associated with the intention to use three out of five PBS and the total MD score. Regression coefficients showed gender invariance for all six models.

**Conclusions:** Our findings suggest that interventions aimed at promoting motivation to use alcohol PBS among young adults could benefit the most from including components that promote self-efficacy to use PBS and the perceived usefulness of PBS for reducing alcohol-related consequences.

**Short Summary:** This study has shown that the coping appraisal components of Protection Motivation Theory have greater explanatory value than threat appraisal components for motivation to use Alcohol Protective Behavioral Strategies related to manner of drinking among young adults who use alcohol. Likewise, the results of this model were invariant across genders.

## Introduction

Alcohol is the most widely consumed substance worldwide, with its use especially prevalent among young adults (World Health Organization [WHO] 2022). Alcohol use — particularly heavy drinking — has been associated with numerous negative consequences, including physical illness, mental health problems, risky sexual behavior, and unsafe driving (WHO 2022), and with an increased likelihood of developing an alcohol use disorder (Prince et al. 2019). Previous research has shown that young adults use Protective Behavioral Strategies (PBS; Martens t al., 2005) defined as cognitive-behavioral strategies to minimize potential alcohol-related consequences. Three subtypes of alcohol PBS have been typically identified: serious harm reduction (SHR) strategies (e.g., eating before or during drinking), stopping/limiting drinking (SLD) strategies (e.g., alternating between alcoholic and nonalcoholic drinks), and manner of drinking (MD) strategies. These latter strategies include avoiding drinking games or mixing different types of alcohol; drinking slowly rather than gulping or chugging; avoiding trying to keep up or out-drink others; and avoiding pregaming. The previous literature has consistently identified these MD strategies as having the highest predictive capacity for reducing intensive alcohol consumption and its associated consequences in comparison to SLD and SHR strategies (e.g., Fernández-Calderón et al. 2021).

Given the demonstrated importance of PBS in reducing alcohol-related negative consequences, in recent years, a variety of studies have analyzed the explanatory factors of their use (for a review, see González-Ponce, Rojas-Tejada et al. 2022), including perceived efficacy of PBS to reduce alcohol-related negative consequences (González-Ponce, Carmona et al. 2022), perceived self-efficacy to engage in PBS (Ray et al. 2009), and descriptive social norms regarding the use of PBS within peer groups (Fernández-Calderón et al. 2022). Although identifying these factors may be useful for designing interventions aimed at promoting the use of PBS, theory-based explanatory models have shown greater utility in explaining the development, maintenance, and change in patterns of health-related behaviors (Conner and Norman, 2015). In addition, health-promotion interventions based on theoretical models are more effective than those based on isolated constructs (e.g., DiClemente et al. 2013). Despite the above, to the best of our knowledge, only one study has attempted to explain the use of PBS based on a theoretical model. In particular, Scaglione et al. (2015) demonstrated the utility of a dual-process decision-making in predicting PBS use among college students.

The Protection Motivation Theory (PMT; Maddux and Rogers, 1983; Rogers 1983) is one of the most widely used theoretical models in the field of health psychology, having shown a high predictive power for intention to engage in various health-protective behaviors, including smoking cessation (Maddux and Rogers, 1983), and condom use (Chambers et al. 2018). This theory was developed to explain the motivation (operationalized as an intention) to engage in health-protective behaviors in the face of a potential health threat. In particular, as posed by the revised theory formulated by Maddux and Rogers (1983), it is expected that the intention to adopt a health-protective behavior depends on two cognitive processes that are triggered when a health threat appears: threat appraisal and coping appraisal. Threat appraisal is determined by the likelihood of experiencing the threat (perceived vulnerability) and the perceived severity of the health threat, while coping appraisal is composed of beliefs about whether a protective behavior (e.g., using PBS) will be effective in reducing the threat (response efficacy) and the belief in one’s ability to execute the coping response successfully (self-efficacy). According to the postulates of PMT, the intention to engage in a protective behavior in the face of a threat will be greater when the individual perceives that the threat is serious and probable (threat appraisal), that they possess the skills to execute such a behavior, and that the protective behavior will be effective in reducing the threat (coping appraisal).

While numerous studies (e.g., Floyd et al., 2000) have yielded support for this model in the context of various health-promoting behaviors, several systematic reviews and meta-analyses (e.g., Ruiter et al. 2014) have shown that the coping appraisal component (perceived self-efficacy and response efficacy) has greater explanatory power than threat appraisal. In addition, intention has typically emerged as the strongest predictor of behavior (Azjen, 1991), being considered as the last step of the motivational phase of decision making (Conner & Norman, 2015). In the field of alcohol use, the coping appraisal component has shown to be relevant in explaining PBS use (e.g., González-Ponce, Carmona-Márquez et al. 2022), and it has been shown that intention to use PBS among university students is associated with PBS use (Fairlie et al., 2021). Thus, knowing the explanatory power of PMT and its various constructs for the intention to use alcohol PBS could help to inform the design of effective interventions to encourage motivation to use these strategies among young adults.

Previous studies have shown that women experience more alcohol-related negative consequences than men (Patrick et al. 2020). Moreover, women’s affective response to threatening stimuli is more negative (Wen et al. 2022), and, therefore, they tend to protect themselves more from the same threat than men (Guo et al. 2015). In coherence, previous research has demonstrated that women use PBS more frequently than men (e.g., Schwebel et al. 2022). This use is associated with more reductions in alcohol-related consequences for women than for men (Clarke et al. 2016). The explanatory factors of PBS use (e.g., perceived vulnerability and perceived severity of the threat) may impact males and females differently and produce higher or lower use of PBS. Thus, examining whether there are gender differences in the explanatory power of the PMT for PBS use could be informative for designing targeted gender-based interventions for promoting the use of these strategies.

Considering the above, we aimed to i) prospectively examine the explanatory value of the PMT for the intention to use alcohol PBS related to the manner of drinking (MD) strategies, and ii) examine whether this explanatory model varies across gender. Based on the previous literature, it is hypothesized that the coping appraisal components will show greater explanatory power than those of threat appraisal. However, given the lack of research that has examined gender differences in the explanatory value of the PMT for PBS use, no hypothesis is proposed in this regard.

## Method

### Participants and Procedure

Participants were young adults aged 18-25 years (*M* = 21.1 [*SD* = 2.2], females = 50.7%) who reported past-month alcohol use on at least two occasions. The participants were recruited through targeted sampling (Watter and Biernacki 1989) from several predetermined community contexts in the province of Huelva (e.g., parks, bars). A psychologist with experience in research was responsible for recruitment and questionnaire administration. In addition, posters were used to recruit participants, and following the targeted sampling protocol (Watter and Biernacki, 1989), snowball sampling was used. Of the total sample (*n* = 360), 48.3% (*n* = 174) of the participants were recruited directly by the fieldworker, 43.1% (*n* = 155) were identified by other participants, and 8.6% (*n* = 31) contacted the researcher after seeing a poster in the street. Participants gave informed consent before taking part and completed the questionnaires in paper and pencil format in designated rooms at the University of Huelva. Each participant was compensated with an Amazon voucher for 15 euros.

From the baseline sample (*n* = 360), 94.2% (*n* = 339) completed a 2-month follow-up. Similar to the baseline measure, participants received an Amazon voucher for 15 euros. Further details of this procedure can be found in Fernández-Calderón et al. (2021).

Among those who participated in the follow-up, 96.2% reported being born in Spain, and 59.0% were studying at a university at the time of the study. The main sources of income reported were a family allowance (51.6%) or a paid job (25.1%), and 77.6% lived with their parents. At baseline, the mean number of drinking and binge drinking days in the past two months was 15.8 (*SD* = 11.5) and 5.7 (*SD* = 7.2), respectively.

No statistically significant differences were found between those who participated in the follow-up survey (*n* = 339) and those who did not (*n* = 21) in terms of gender, age, and mean days of drunkenness in the past-2 months (all *p* > 0.05). However, significant differences were found regarding the mean days of alcohol use in the past-2 months (Mann-Whitney *U* = 2628; *z* = -2.016, *p* = .044). It was found that those who participated in the follow-up reported more days of consumption (*M* = 15.79; *SD* = 11.54) than those who did not (*M* = 10.62; *SD* = 7.19).

The present study was approved by the Regional Bioethics Research Committee of Andalusia (Consejería de Sanidad, Junta de Andalucía, Spain).

### Measures

The following measures were included in the final questionnaire:

*Alcohol use measures (baseline):* Participants reported the number of days of alcohol consumption and the number of days they were drunk in the last two months.

#### Perceived vulnerability to alcohol consequences (baseline)

Following previous studies (e.g., Vera et al. 2022), two items were used to measure perceived vulnerability to experiencing alcohol-related consequences. Participants were asked to indicate how likely they were to experience negative health consequences from drinking alcohol (Item 1) and getting drunk (Item 2) in a five-point response format (1-very unlikely to 5-very likely). Responses were summed to obtain a total score of perceived vulnerability. The Spearman-Brown reliability coefficient for perceived vulnerability was .82.

#### Perceived severity of alcohol consequences (baseline)

Perceived severity was assessed in a similar way to perceived vulnerability. Thus, following previous research (e.g., Vera et al. 2022), two items were used to ask participants how risky it is to consume alcohol and get drunk (ordinal response format ranging from 1-very unlikely to 5-very likely). The total score was obtained by summing the responses to the two items. The Spearman-Brown reliability coefficient for perceived severity was .73.

#### Perceived Efficacy of MD PBS to reduce alcohol-related negative consequences (baseline)

Similar to previous studies (e.g., Ray et al. 2009), we used a modified version of the MD subscale of the PBSS (Treloar et al. 2015) in its Spanish version (Sánchez-García et al. 2020) to measure perceived efficacy of the five PBS. Participants were asked: “*Please, indicate how effective each of the following behaviors are in reducing alcohol-related negative consequences.”* We used the same response options as Ray et al. (2009; 1-not at all effective, 2-somewhat effective, 3-moderately effective, 4-extremely effective). Cronbach’s alpha of the perceived efficacy MD scale was .77.

#### Perceived Self-efficacy to engage in MD PBS (baseline)

Based on the recommendations by Ajzen (2006) and previous research in the field of PBS use (Davis and Rosenberg 2016), three items were created to evaluate perceived self-efficacy to use each of the five MD PBS. For example, for the strategy *Avoid mixing different types of alcohol*, participants were asked about the extent to which they agree (from 1 = totally disagree to 7 = totally agree) with the following statements: “If I wanted to, I would be able to avoid mixing different types of alcohol,” “*if I choose to avoid mixing different types of alcohol when I drink, I can do so,*” and “*If I wanted to, I could easily avoid mixing different types of alcohol.”* The Cronbach alpha values were as follows: self-efficacy to avoid drinking games =. 89, avoiding mixing =. 79, drinking slowly =. 91, avoiding keeping up =.87, avoiding pregaming =. 90, and the total score on the MD self-efficacy scale =. 93.

#### Intention to use MD PBS (at follow-up)

As in the case of self-efficacy measurements, we considered Ajzen’s recommendations (2006) and previous research (Davis and Rosenberg 2016) to assess the intention to use each MD PBS. For example, for the behavior Avoid mixing different types of alcohol, participants were asked to indicate their degree of agreement (from 1 = totally disagree to 7 = totally agree) with the following statements: *“In the next two months, I am likely to avoid mixing different types of alcohol’, ’In the next two months, when I drink I intend to avoid mixing different types of alcohol’* and *’In the next two months, I will avoid mixing different types of alcohol.’* Ordinal Cronbach’s alpha values were as follows: the intention to avoid drinking games=. 98, avoiding mixing = .93, drinking slowly = .96, avoiding keeping up = .94, avoiding pregaming =. 96, and total score on intention to use MD PBS scale =. 95.

### Data analysis

To address the research questions, we used SPSS 26 (IMB, 2016) to conduct six multiple linear regression models, one for each of the five behavior intentions and one for the total score on the MD intention scale as the outcome variables. These models allowed us to estimate the potential associations between PMT variables (perceived vulnerability, perceived severity, response efficacy, and self-efficacy) at baseline and the intentions to use MD PBS at follow-up. Participants’ age and gender, alcohol use measures (frequency of alcohol and drunkenness) and intention to use MD PBS at baseline were included as control variables in all the regression models.

Mplus 8.7 (Muthén and Muthén 2017) multigroup regression analyses were used to evaluate the invariance of the models across gender Multigroup models were specified with all the regression coefficients restricted to be equal for men and women. Three indices were used to evaluate the fit of the model: root mean square error of approximation (RMSEA), comparative fit index (CFI), and standardized root mean squared residual (SRMR). Accepted CFI values are >0.95, and the RMSEA and SRMR should be <0.08. The fit of the constrained models was considered a measure of the plausibility of the invariance hypotheses.

## Results

### Descriptive and Bivariate Results

Means, standard deviations, and bivariate correlations between predictors and outcome variables are presented in Table 1. The baseline perceived severity score was positively related to the intention to use all behaviors except for ‘Avoiding pregaming’. In contrast, the baseline perceived vulnerability score was not associated with intention to use any of the behaviors. Regarding coping components, perceived efficacy and self-efficacy at baseline were positively associated with the intention to use PBS at follow-up.

**Table 1.** Descriptive statistics and correlations between study variables.

|  |  | Intention<br>ADG | Intention<br>AM | Intention<br>DS | Intention<br>AKU | Intention<br>AP | Intention<br>TM |
| --- | --- | --- | --- | --- | --- | --- | --- |
|  | *M (SD)* | 11.77 (5.90) | 15.84 (5.18) | 15.27 (4.96) | 16.04 (4.71) | 12.39 (5.45) | 71.60 (20.46) |
| Age | 21.15 (2.23) | .144<sup>**</sup> | .145<sup>**</sup> | .154<sup>**</sup> | .129<sup>*</sup> | .129<sup>*</sup> | .188<sup>**</sup> |
| Alcohol use | 15.49 (11.39) | -.206<sup>**</sup> | -.254<sup>**</sup> | -.295<sup>**</sup> | -.201<sup>**</sup> | -.285<sup>**</sup> | -.301<sup>**</sup> |
| Drunkenness | 5.70 (7.11) | -.277<sup>**</sup> | -.301<sup>**</sup> | -.393<sup>**</sup> | -.284<sup>**</sup> | -.321<sup>**</sup> | -.393<sup>**</sup> |
| Perceived severity | 8.25 (1.35) | .122<sup>*</sup> | .171<sup>**</sup> | .252<sup>**</sup> | .236<sup>**</sup> | .067 | .202<sup>**</sup> |
| Perceived vulnerability | 6.29 (1.98) | .042 | .013 | -.047 | -.009 | -.022 | -.021 |
| Perceived efficacy of ADG | 3.02 (0.88) | .306<sup>**</sup> | .156<sup>**</sup> | .225<sup>**</sup> | .171<sup>**</sup> | .219<sup>**</sup> | .282<sup>**</sup> |
| Perceived efficacy of AM | 3.60 (0.70) | .101 | .307<sup>**</sup> | .241<sup>**</sup> | .196<sup>**</sup> | .091 | .241<sup>**</sup> |
| Perceived efficacy of DS | 3.54 (0.67) | .108<sup>*</sup> | .229<sup>**</sup> | .315<sup>**</sup> | .281<sup>**</sup> | .171<sup>**</sup> | .282<sup>**</sup> |
| Perceived efficacy of AKU | 3.39 (0.82) | .115<sup>*</sup> | .164<sup>**</sup> | .233<sup>**</sup> | .252<sup>**</sup> | .102 | .213<sup>**</sup> |
| Perceived efficacy of AP | 2.87 (0.87) | .122<sup>*</sup> | .181<sup>**</sup> | .115<sup>*</sup> | .079 | .294<sup>**</sup> | .202<sup>**</sup> |
| Perceived efficacy of TM | 16.43 (2.60) | .240<sup>**</sup> | .305<sup>**</sup> | .330<sup>**</sup> | .293<sup>**</sup> | .277<sup>**</sup> | .371<sup>**</sup> |
| Perceived Self-efficacy of ADG | 18.74 (3.32) | .185<sup>**</sup> | .187<sup>**</sup> | .241<sup>**</sup> | .221<sup>**</sup> | .104 | .245<sup>**</sup> |
| Perceived Self-efficacy of AM | 19.32 (2.53) | .062 | .227<sup>**</sup> | .234<sup>**</sup> | .243<sup>**</sup> | .073 | .212<sup>**</sup> |
| Perceived Self-efficacy of DS | 18.64 (3.34) | .107 | .205<sup>**</sup> | .407<sup>**</sup> | .302<sup>**</sup> | .134<sup>*</sup> | .303<sup>**</sup> |
| Perceived Self-efficacy of AKU | 18.86 (2.98) | .086 | .211<sup>**</sup> | .304<sup>**</sup> | .333<sup>**</sup> | .133<sup>*</sup> | .274<sup>**</sup> |
| Perceived Self-efficacy of AP | 18.86 (3.59) | .088 | .076 | .193<sup>**</sup> | .211<sup>**</sup> | .232<sup>**</sup> | .205<sup>**</sup> |
| Perceived Self-efficacy of TM | 94.39 (11.39) | .150<sup>**</sup> | .251<sup>**</sup> | .387<sup>**</sup> | .375<sup>**</sup> | .197<sup>**</sup> | .351<sup>**</sup> |

*Note.* ADG = “avoiding drinking games”, AM = “avoiding mixing different types of alcohol”, DS = “drinking slowly”, AKU = “avoiding keeping up” AP = “avoiding pregameing”, TM= Total score in the MD subscale \*p<.05, \*\*p<.01.

### Regression Results

The results of the multiple regression predicting intention to use MD PBS as a function PMT variables are shown in Table 2. After controlling for the effects of sociodemographic variables, alcohol use, and intention to use MD PBS at baseline, the variance explained by the PMT constructs ranged from 8% for the intention to avoid drinking games to 16% for the intention to drink slowly. Perceived efficacy and perceived self-efficacy were identified as significant predictors of the intention outcome in all the regression models (Table 2). On the other hand, while perceived vulnerability was not a significant predictor of any of the intention outcomes, perceived severity was associated with the intention to use three out of five MD PBS. All the significant coefficients indicated a positive association between the PMT predictors and the intentions to engage in MD strategies.

**Table 2.**
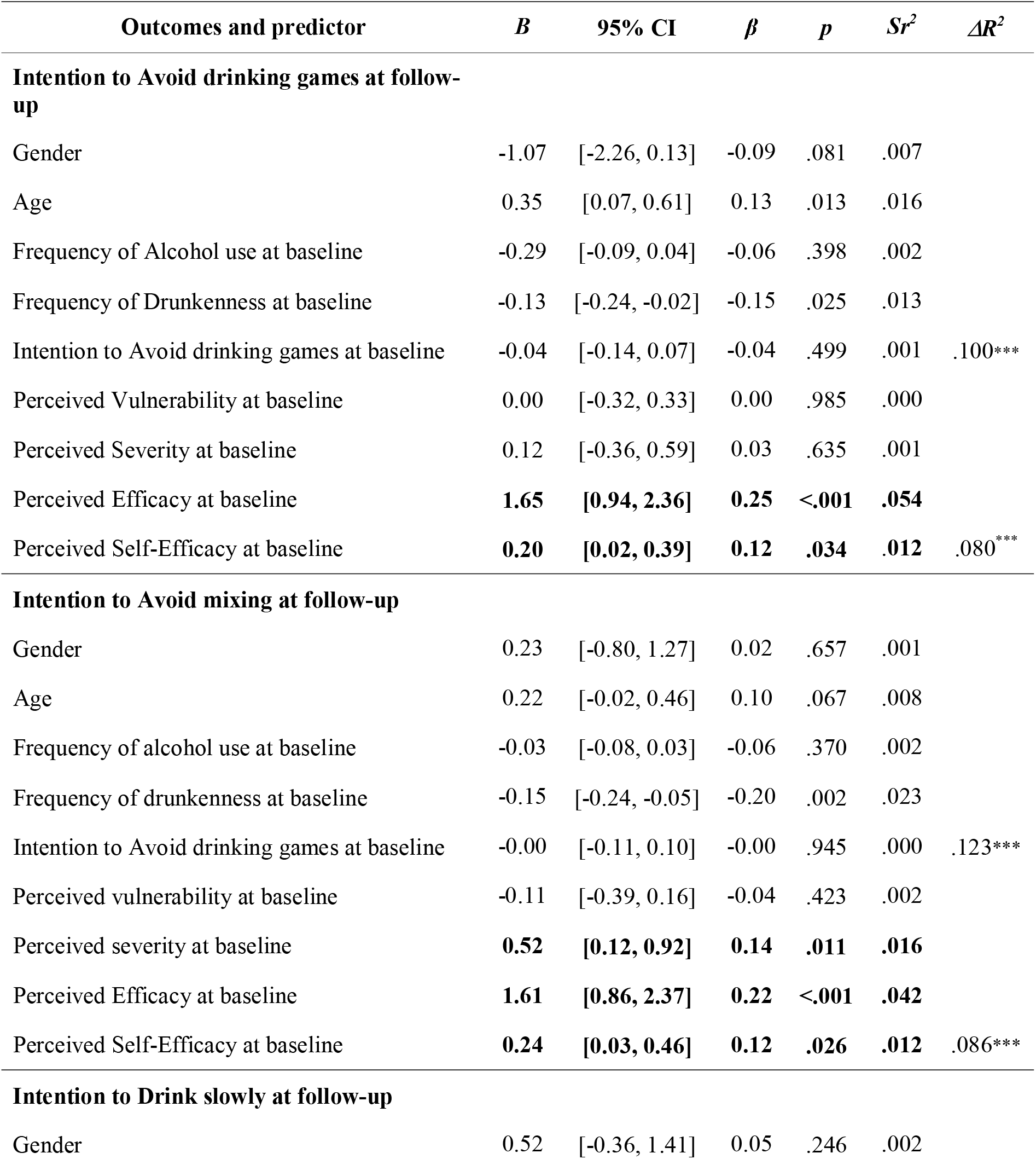
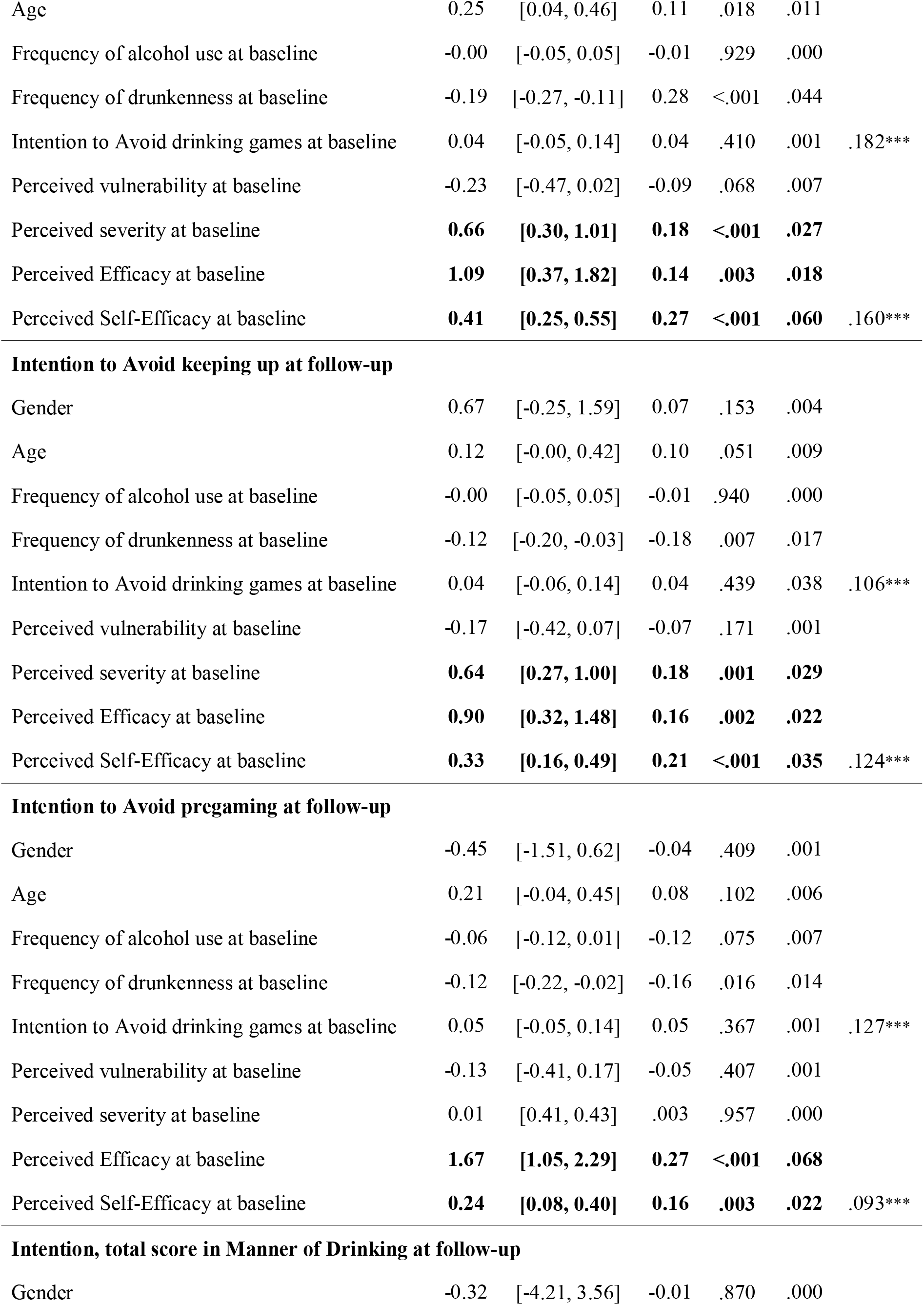
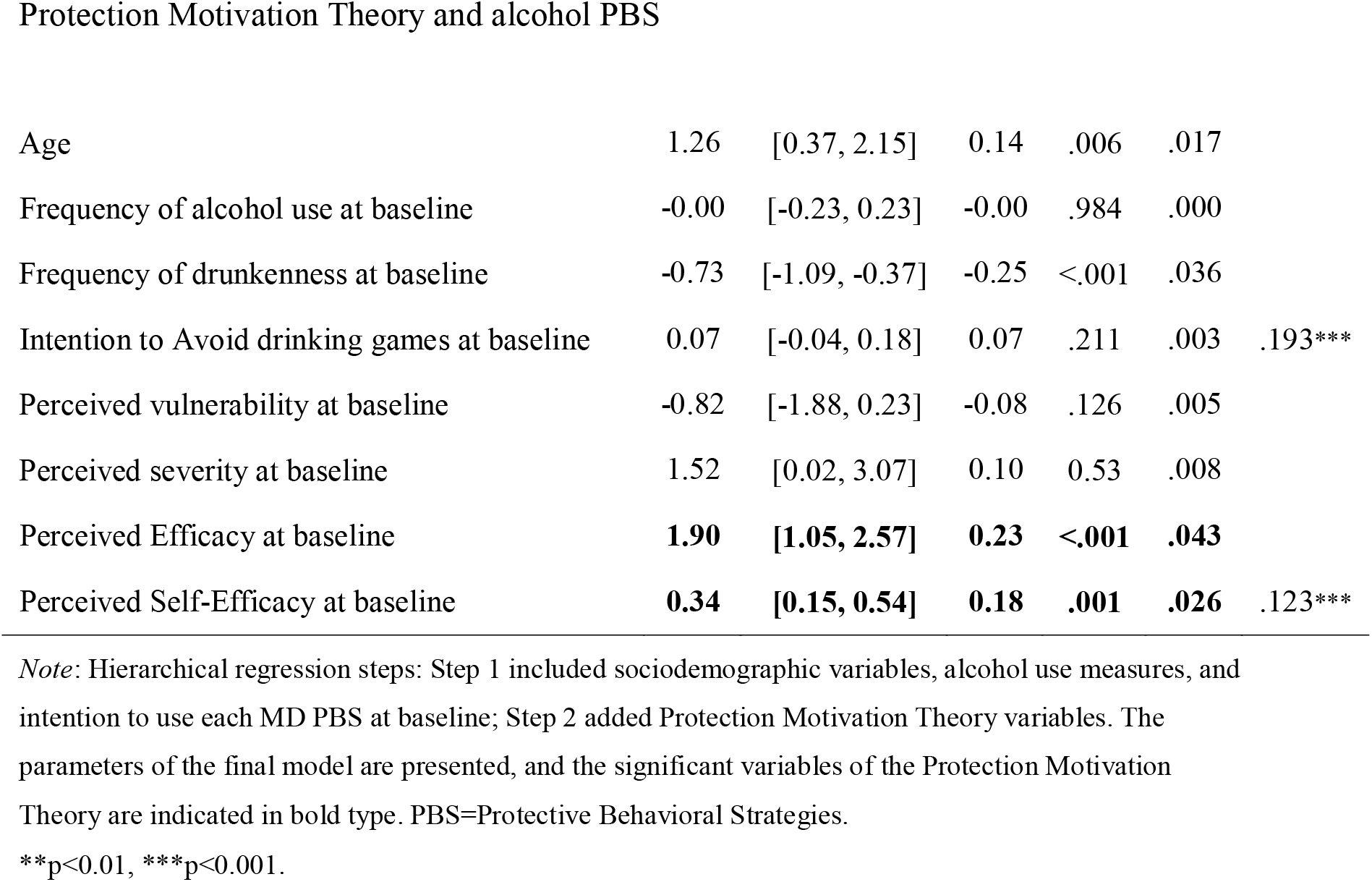
Hierarchical multiple regression examining the association between Protection Motivation Theory variables at baseline and the intentions to use manner of drinking (MD) PBS at follow-up, controlling for sociodemographic variables, alcohol use measures, and intention of the use MD PBS.

| Outcomes and predictor | *B* | 95% CI | $\beta$ | *p* | *Sr*<sup>2</sup> | $\Delta R^2$ |
| --- | --- | --- | --- | --- | --- | --- |
| **Intention to Avoid drinking games at follow-up** |  |  |  |  |  |  |
| Gender | -1.07 | [-2.26, 0.13] | -0.09 | .081 | .007 |  |
| Age | 0.35 | [0.07, 0.61] | 0.13 | .013 | .016 |  |
| Frequency of Alcohol use at baseline | -0.29 | [-0.09, 0.04] | -0.06 | .398 | .002 |  |
| Frequency of Drunkenness at baseline | -0.13 | [-0.24, -0.02] | -0.15 | .025 | .013 |  |
| Intention to Avoid drinking games at baseline | -0.04 | [-0.14, 0.07] | -0.04 | .499 | .001 | .100*** |
| Perceived Vulnerability at baseline | 0.00 | [-0.32, 0.33] | 0.00 | .985 | .000 |  |
| Perceived Severity at baseline | 0.12 | [-0.36, 0.59] | 0.03 | .635 | .001 |  |
| Perceived Efficacy at baseline | **1.65** | **[0.94, 2.36]** | **0.25** | **<.001** | **.054** |  |
| Perceived Self-Efficacy at baseline | **0.20** | **[0.02, 0.39]** | **0.12** | **.034** | **.012** | .080*** |
| **Intention to Avoid mixing at follow-up** |  |  |  |  |  |  |
| Gender | 0.23 | [-0.80, 1.27] | 0.02 | .657 | .001 |  |
| Age | 0.22 | [-0.02, 0.46] | 0.10 | .067 | .008 |  |
| Frequency of alcohol use at baseline | -0.03 | [-0.08, 0.03] | -0.06 | .370 | .002 |  |
| Frequency of drunkenness at baseline | -0.15 | [-0.24, -0.05] | -0.20 | .002 | .023 |  |
| Intention to Avoid drinking games at baseline | -0.00 | [-0.11, 0.10] | -0.00 | .945 | .000 | .123*** |
| Perceived vulnerability at baseline | -0.11 | [-0.39, 0.16] | -0.04 | .423 | .002 |  |
| Perceived severity at baseline | **0.52** | **[0.12, 0.92]** | **0.14** | **.011** | **.016** |  |
| Perceived Efficacy at baseline | **1.61** | **[0.86, 2.37]** | **0.22** | **<.001** | **.042** |  |
| Perceived Self-Efficacy at baseline | **0.24** | **[0.03, 0.46]** | **0.12** | **.026** | **.012** | .086*** |
| **Intention to Drink slowly at follow-up** |  |  |  |  |  |  |
| Gender | 0.52 | [-0.36, 1.41] | 0.05 | .246 | .002 |  |
| Age | 0.25 | [0.04, 0.46] | 0.11 | .018 | .011 |  |
| Frequency of alcohol use at baseline | -0.00 | [-0.05, 0.05] | -0.01 | .929 | .000 |  |
| Frequency of drunkenness at baseline | -0.19 | [-0.27, -0.11] | 0.28 | <.001 | .044 |  |
| Intention to Avoid drinking games at baseline | 0.04 | [-0.05, 0.14] | 0.04 | .410 | .001 | .182*** |
| Perceived vulnerability at baseline | -0.23 | [-0.47, 0.02] | -0.09 | .068 | .007 |  |
| Perceived severity at baseline | **0.66** | **[0.30, 1.01]** | **0.18** | **<.001** | **.027** |  |
| Perceived Efficacy at baseline | **1.09** | **[0.37, 1.82]** | **0.14** | **.003** | **.018** |  |
| Perceived Self-Efficacy at baseline | **0.41** | **[0.25, 0.55]** | **0.27** | **<.001** | **.060** | .160*** |
| **Intention to Avoid keeping up at follow-up** |  |  |  |  |  |  |
| Gender | 0.67 | [-0.25, 1.59] | 0.07 | .153 | .004 |  |
| Age | 0.12 | [-0.00, 0.42] | 0.10 | .051 | .009 |  |
| Frequency of alcohol use at baseline | -0.00 | [-0.05, 0.05] | -0.01 | .940 | .000 |  |
| Frequency of drunkenness at baseline | -0.12 | [-0.20, -0.03] | -0.18 | .007 | .017 |  |
| Intention to Avoid drinking games at baseline | 0.04 | [-0.06, 0.14] | 0.04 | .439 | .038 | .106*** |
| Perceived vulnerability at baseline | -0.17 | [-0.42, 0.07] | -0.07 | .171 | .001 |  |
| Perceived severity at baseline | **0.64** | **[0.27, 1.00]** | **0.18** | **.001** | **.029** |  |
| Perceived Efficacy at baseline | **0.90** | **[0.32, 1.48]** | **0.16** | **.002** | **.022** |  |
| Perceived Self-Efficacy at baseline | **0.33** | **[0.16, 0.49]** | **0.21** | **<.001** | **.035** | .124*** |
| **Intention to Avoid pregameing at follow-up** |  |  |  |  |  |  |
| Gender | -0.45 | [-1.51, 0.62] | -0.04 | .409 | .001 |  |
| Age | 0.21 | [-0.04, 0.45] | 0.08 | .102 | .006 |  |
| Frequency of alcohol use at baseline | -0.06 | [-0.12, 0.01] | -0.12 | .075 | .007 |  |
| Frequency of drunkenness at baseline | -0.12 | [-0.22, -0.02] | -0.16 | .016 | .014 |  |
| Intention to Avoid drinking games at baseline | 0.05 | [-0.05, 0.14] | 0.05 | .367 | .001 | .127*** |
| Perceived vulnerability at baseline | -0.13 | [-0.41, 0.17] | -0.05 | .407 | .001 |  |
| Perceived severity at baseline | 0.01 | [0.41, 0.43] | .003 | .957 | .000 |  |
| Perceived Efficacy at baseline | **1.67** | **[1.05, 2.29]** | **0.27** | **<.001** | **.068** |  |
| Perceived Self-Efficacy at baseline | **0.24** | **[0.08, 0.40]** | **0.16** | **.003** | **.022** | .093*** |
| **Intention, total score in Manner of Drinking at follow-up** |  |  |  |  |  |  |
| Gender | -0.32 | [-4.21, 3.56] | -0.01 | .870 | .000 |  |
| Age | 1.26 | [0.37, 2.15] | 0.14 | .006 | .017 |  |
| Frequency of alcohol use at baseline | -0.00 | [-0.23, 0.23] | -0.00 | .984 | .000 |  |
| Frequency of drunkenness at baseline | -0.73 | [-1.09, -0.37] | -0.25 | <.001 | .036 |  |
| Intention to Avoid drinking games at baseline | 0.07 | [-0.04, 0.18] | 0.07 | .211 | .003 | .193*** |
| Perceived vulnerability at baseline | -0.82 | [-1.88, 0.23] | -0.08 | .126 | .005 |  |
| Perceived severity at baseline | 1.52 | [0.02, 3.07] | 0.10 | 0.53 | .008 |  |
| Perceived Efficacy at baseline | **1.90** | **[1.05, 2.57]** | **0.23** | **<.001** | **.043** |  |
| Perceived Self-Efficacy at baseline | **0.34** | **[0.15, 0.54]** | **0.18** | **.001** | **.026** | .123*** |

*Note:* Hierarchical regression steps: Step 1 included sociodemographic variables, alcohol use measures, and intention to use each MD PBS at baseline; Step 2 added Protection Motivation Theory variables. The parameters of the final model are presented, and the significant variables of the Protection Motivation Theory are indicated in bold type. PBS=Protective Behavioral Strategies.
\*\*p<0.01, \*\*\*p<0.001.

The fit of the six multigroup gender-invariant models is presented in Table 3. None of the model chi-squares were significant, indicating that regression coefficients were gender invariant for the six models.

**Table 3.** Multigroup gender-invariant regression models.

|  | *Chi-square (df)* | *p* | *RMSEA* | *CFI* | *SRMR* |
| --- | --- | --- | --- | --- | --- |
| Avoiding drinking games | 2.39 (8) | .967 | 0 | 1 | .023 |
| Avoiding mixing | 2.52 (8) | .961 | 0 | 1 | .025 |
| Drinking slowly | 6.28 (8) | .616 | 0 | 1 | .036 |
| Avoiding keeping-up | 7.75 (8) | .458 | .0 | 1 | .040 |
| Avoiding pregaming | 6.70 (8) | .569 | 0 | 1 | .043 |
| Total MD score | 5.54 (8) | .699 | 0 | 1 | .035 |

*Note.* *df* = degrees of freedom; *p* = p value; *RMSEA* = Root Mean-Square Error of Approximation; *CFI* = Comparative Fit Index; *SRMR* = Standardized Root Mean-Square Residual; MD = manner of drinking.

## Discussion

Theory-based interventions have shown utility in promoting various health-related behaviors (e.g., DiClemente et al. 2013). However, although previous research has examined the psychological determinants of alcohol PBS use (González-Ponce, Rojas- Tejada et al. 2022), theory-based research is scarce. To our knowledge, this is the first study that prospectively examines the explanatory value of one of the most prominent theories in the field of health-related behavior, the Protection Motivation Theory (Maddux and Rogers 1983), for the intention to use alcohol PBS among young adults.

Our findings support the utility of this theory when applied to intention to use PBS and also indicate that the explanatory value of his theory does not differ according to gender.

As hypothesized and consistently shown in previous studies and systematic reviews (e.g., Ruiter et al. 2014), our findings support the greater explanatory value of coping appraisal (response efficacy and self-efficacy) in comparison to threat appraisal (vulnerability and severity). These findings are in line with previous studies showing the association between coping appraisal components (response efficacy and self-efficacy) and PBS use (González-Ponce, Carmona-Márquez et al. 2022). Also, this finding supports the assertion made by Ruiter et al. (2014) following their meta-analytic review, who highlighted the low impact of presenting threatening health-related information (i.e., fear appeal) when promoting protective behaviors. In contrast, promoting PBS efficacy to reduce alcohol-related negative consequences, and increasing perceived self-efficacy to use PBS, appear to be more effective ways of increasing the motivation to use alcohol PBS in young adults.

Regarding the threat appraisal components, previous meta-analytic reviews have shown a higher utility of perceived severity in explaining health-related behaviors (e.g., Bui et al. 2013), while others point out perceived vulnerability as the most explanatory component of threat appraisal (e.g., Milne et al. 2000). Our results suggest the importance of the former since no prospective association was found between perceived vulnerability and PBS intentions. However, perceived severity was associated with the intention to use three out of five PBS and with the total MOD PBS score, which suggests that providing information about the severity of alcohol’s negative consequences may help to encourage alcohol-using young adults to use PBS.

Regarding coping appraisal, in line with previous research (e.g., Ruiter et al. 2014), beliefs about response efficacy and the ability to engage in PBS may be the best predictors of the intentions to use PBS. Previous meta-analytic reviews (e.g., Bui et al. 2013) have shown self-efficacy to be a stronger predictor than response efficacy. These findings are consistent with the relevance of self-efficacy as a core construct in other prominent theories in the field of human behavior, such as the theory of planned behavior (Ajzen 1991) and social cognitive theory (Bandura 1977). Our findings do not support this superiority of self-efficacy over response efficacy since the association between baseline perceived self-efficacy and follow-up intention was stronger than that between response efficacy and intention for only two out of the five PBS (‘Drinking slowly’ and ‘Avoiding keeping up or out-drinking others’). Instead, both coping components seem to have a comparable impact on intention to use alcohol PBS.

As consistently shown in previous research (e.g., Schwebel et al. 2022), women use more PBS than men, which may be explained by the fact that when faced with a health threat, the affective response of women is higher than that of men. However, our results showed that the relationships between PMT constructs and intention to use the five alcohol PBS examined were invariant across genders. This finding is in line with the results of Plotnikooff et al. (2009), who found gender invariance when examining the explanatory value of the PMT in the field of physical activity, suggesting that interventions based on the PMT may have a similar impact on both men and women.

## Limitations and Future Directions

Some limitations should be considered when interpreting our results. First, previous research (e.g., Witte and Allen et al. 2000) has found that fear appeal seems to be more effective when threats are explicitly described by giving examples of particular negative consequences (e.g., loss of consciousness, vomiting) instead of referring to a broad category of negative consequences. However, in our study, we asked about the perceived vulnerability and severity of health consequences of drinking and binge drinking but did not provide concrete examples of such threats. This aspect could explain why we observed a lower explanatory power of threat appraisal than coping appraisal.

Moreover, when interpreting our results, it should be considered that we did not use a probabilistic sampling procedure, which limits our ability to interpret our findings as being representative of the wider alcohol-using young adult population. Finally, it should be noted that, in line with the tenets of PMT, we examined the explanatory value of this theoretical model for the motivation (i.e., intention) to use alcohol PBS. However, we did not include a second follow-up survey to collect information about actual PBS use. In this regard, it is well known (Conner and Norman 2022) that behavioral intention only partially explains future behavior (the intention-behavior gap). Thus, future studies should include a follow-up measure of behavior to determine the extent to which intention to use PBS translates to actual use of these strategies in the short term. In addition, it would be of interest to include representative samples of young adults and more specific measures of potential alcohol-related consequences.

## Conclusions

Our findings demonstrate the utility of the Protection Motivation Theory in explaining the intention to use a set of alcohol Manner of Drinking PBS, which are associated with less intensive alcohol consumption and fewer negative consequences. In particular, our findings suggest that interventions aimed at promoting intention to use alcohol PBS among young adults could particularly benefit from including components that promote self-efficacy to use PBS and the perceived usefulness of such strategies.

## Data availability

The data underlying this article will be shared on reasonable request to the corresponding author [FFC].

**CRediT: Bella M. González-Ponce**: Conceptualization; Data curation; Investigation; Methodology; Writing-original draft; Data curation; Writing - review & editing. **José Carmona-Márquez**: Data curation; formal analysis; Supervision. **Angelina Pilatti**: Methodology; Writing-review & editing. **Carmen Díaz-Batanero**: Methodology; Writing-review & editing. **Fermín Fernández-Calderón**: Conceptualization; Data curation; Investigation; Methodology; Writing-review & editing; Supervision, Project administration; Funding acquisition.

